# Characterization of the Second Wave of COVID-19 in India

**DOI:** 10.1101/2021.04.17.21255665

**Authors:** Rajesh Ranjan, Aryan Sharma, Mahendra K. Verma

## Abstract

The second wave of COVID-19, which began around 11 February 2021, has hit India very hard with the daily cases reaching nearly triple the first peak value as on April 19, 2021. The epidemic evolution in India is quite complex due to regional inhomogeneities and the spread of several coronavirus mutants. In this paper, we characterize the virus spread in the ongoing second wave in India and its states until April 19, 2021, and also study the dynamical evolution of the epidemic from the beginning of the outbreak. Variations in the effective reproduction number (*R*_*t*_) are taken as quantifiable measures of the virus transmissibility. *R*_*t*_ value for every state, including those with large rural populations, has value greater than the self-sustaining threshold of 1. An exponential fit on recent data also shows that the infection rate is much higher than the first wave. Subsequently, characteristics of the COVID-19 spread are analyzed regionwise, by estimating test positivity rates (TPRs) and case fatality rates (CFRs). Very high TPR values for several states present an alarming situation. CFR values are lower than those in the first wave but recently showing signs of increase as healthcare systems become over-stretched with the surge in infections. Preliminary estimates with a classical epidemiological model suggest that the peak for the second wave could occur around mid-May 2021 with daily count exceeding 0.4 million. The study strongly suggests that an effective administrative intervention is needed to arrest the rapid growth of the epidemic.

## 1 Introduction

More than one year since Coronavirus Disease 2019 (COVID-19) was declared a pandemic on March 11, 2020, by World Health Organization (WHO), the deadly SARS-CoV-2 virus continues to disrupt public life across the world. Although the lockdown norms are relatively relaxed in most places, social life is still far from normal. Recently, multiple vaccines developed by Oxford–AstraZeneca (Covishield/Vaxzevria), Pfizer–BioNTech (Comirnaty), Moderna, Johnson & Johnson’s Janssen, Bharat Biotech (Covaxin), Gamaleya Research Institute of Epidemiology and Microbiology (Sputnik V) etc. have been approved in several countries and are given on priority to susceptible populations and those with co-morbidities. However, the production and distribution of vaccines at a massive scale to cover a very large population remain a formidable challenge. Meanwhile, in order to arrest the spread of the virus during the vaccination drive, intervention measures such as wearing masks, social distancing guidelines, partial lockdowns, and restricted store hours are still in place in most places.

While nations are taking extensive measures to accelerate the vaccination drive in order to control the pandemic at the earliest, a public health challenge has appeared due to mutations of the SARS-CoV-2 virus which make it highly contagious. For example, the SARS-CoV-2 lineage B.1.1.7, which was first detected in the United Kingdom (UK) in November 2020, is estimated to be 40-80% more transmissible than the wild type SARS-CoV-2 [1, 2]. Similarly, strains from South Africa (B.1.351), Brazil (P.1), and India (B.1.617) are also significantly more contagious than the variants in early 2020 [3]. There is no clear evidence on the severity of the new mutations [1], however the challenge is to prepare for health response especially when the number of infections is exceedingly large.

In order to understand the impact of spread of mutated coronavirus, we show the temporal variations of daily COVID-19 cases and deaths in the top six countries in figures 1(a) and (b) respectively. These countries are most affected in terms of the maximum number of cumulative cases as of April 19, 2021. Among the countries shown, the United States (US) has maximum cumulative cases with a total of about 32.5 million, followed by India that has 15.6 million cases. Although if we compare the recent epidemic growth in these countries as shown in the figure, the daily number of infections in India is several times than that in the US. The plot also shows the peaks of the first and second waves in different countries where they were observed. The US, for example, had the first peak in mid-July 2020 following which the cases subsided. However, the infection counts started increasing again in October and a much larger peak was observed in the second wave in December 2020 with daily cases of up to 0.25 million. In the UK, the first and second peaks were separated by only a couple of months with the second wave largely attributed to a more infectious mutant [4]. In both countries, the peak of the second wave is much higher than the first wave. For Russia, the infection curve is relatively shallower with the number of daily cases never crossing 30,000. In the curve for Brazil, there are large fluctuations which may be due to insufficient testing, however, the number of deaths per infection (typically called *Case Fatality Rate* (*CFR*)) is very large, as can be inferred from figure 1(b). The curve for the death cases typically follows the infections. Note, however, that the CFR was very high at the beginning of the pandemic (around April 2020) for the US and European countries [5].

**Figure 1:**
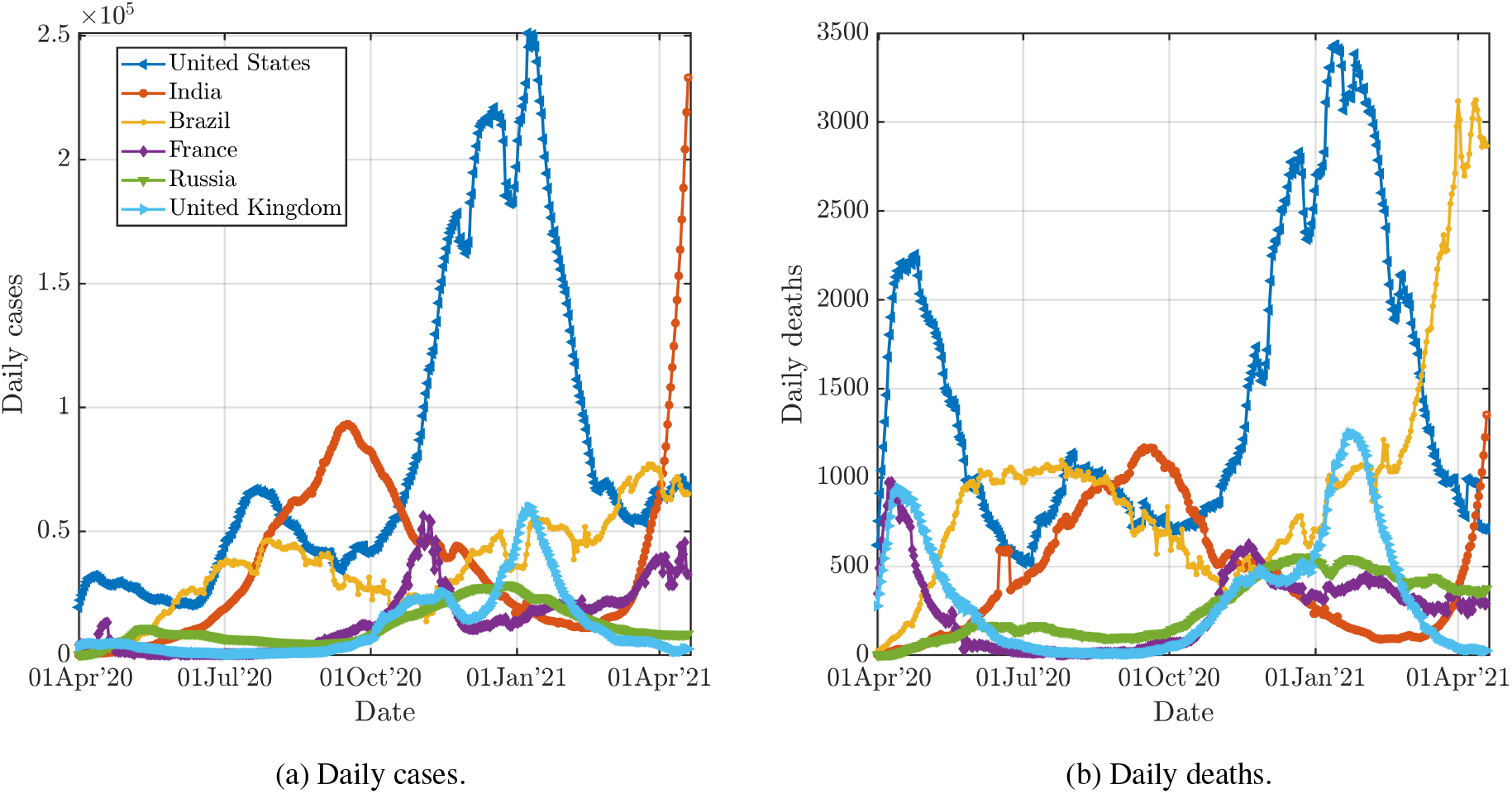
Comparison of COVID-19 spread in India with five other severely-affected countries. Smoothened data between April 1, 2020 and April 19, 2021 as retrieved from OWID are used for the plots. Variations in daily number of new infections (a) and new deaths (b) are shown. Curves clearly show the first and second waves for the US, UK, and India. Peaks in second waves for all these countries are higher than those in first waves.

For India, as seen in figures 1(a) and (b), the first and second waves are separated by about 5 months. The peak of the first wave was in September 2020 with daily cases of around 0.1 million. The daily cases decreased until mid-February after which it exhibited a sharp increase. The end of the first wave was likely a result of a combination of factors – effective implementation of government interventions, increase in awareness, and, most importantly, the experience gained by medical professionals in treating the disease over the initial months. On April 19, 2021, the number of new cases was about 0.3 million which is already triple the first peak value. The sudden surge in the number of cases after a relatively long ‘cooling’ time is baffling although it may be attributed to a highly infectious double mutant variant of SARS-CoV-2 (B.1.617 lineage), to the complacent behaviour of the population, and to the relaxation of interventions [6]. The number of daily deaths is also rising recently, but the CFR is low compared to the first wave; this aspect will be further discussed later. Note that the study on B.1.617 mutant is limited and there is no clear evidence on whether the mutant virus is less severe than its predecessor.

The present study focuses on the ongoing second wave in India and therefore, we look at the regional distribution of COVID-19 spread. Figures 2(a) and (b) show the daily numbers of reported infections in 16 key states in India, in linear and log scales respectively. In figure 2(a), we note that almost all the states are showing a surge in the number of cases since 11 February, 2021. Further, the slopes of the growth curves are very steep in the second wave compared to the first. The daily number of cases in Maharashtra, which also leads in the daily as well as cumulative infections, went from daily cases of 652 on February 11, 2021 to about 63,000 in two months (as on April 11, 2021). This rapid growth is also observed in other states (see figure 2(b)), albeit the number of daily cases is fewer than that in Maharashtra as on April 19, 2021. The growth curve in the second wave can be further divided into relatively slow and fast growth phases as shown by green and red shadows in figure 2(a,b). In the first region, until the first week of March, all states except Maharashtra exhibited a slow increase in the number of cases. However, in the second region, most states show a sudden spurt in the number of infections propelling India’s total daily count to about 0.3 million.

**Figure 2:**
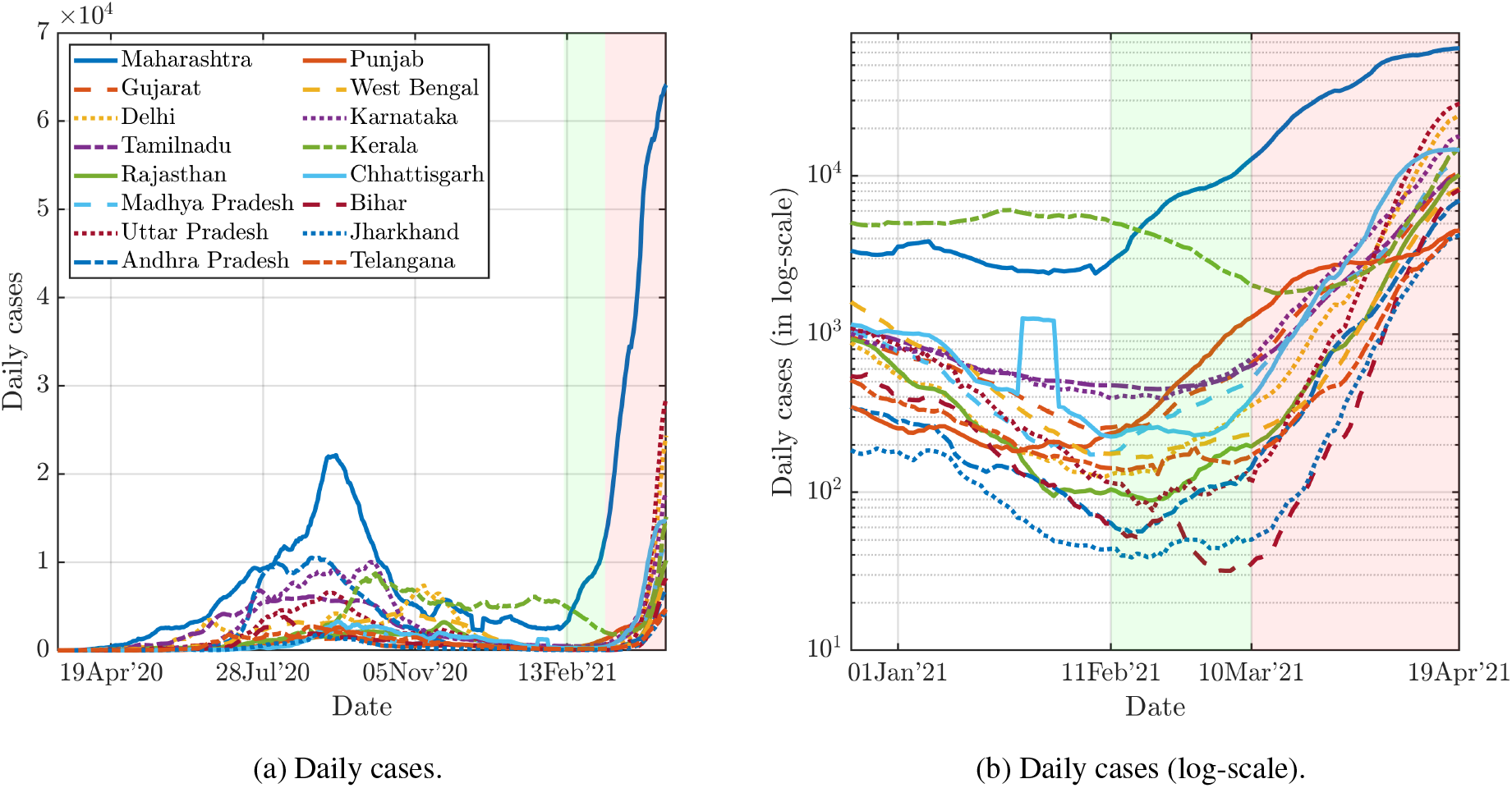
Comparison of COVID-19 spread in most-impacted 16 states of India as on April, 19, 2021. Time-series infection data are taken from covid19india.org and are smoothened by taking 7-day averaging. Figure (a) shows the variations in daily infections from the beginning of the pandemic, while figure (b) shows a zoomed view focusing on the second wave. Green and orange shadows relatively slow and subsequent fast growth of daily cases in the second wave. The top three states in terms of new infections are Maharashtra, Uttar Pradesh, and Delhi.

At the outset, the second wave in India looks much more precarious than the first wave. Although India is the fastest in the world to administer COVID-19 vaccine doses [7], the fraction of the vaccinated population is relatively small due to the very large population of the country. Further, India is yet to administer vaccines to populations less than 45 years of age (except susceptible populations like doctors, frontline workers), many of whom are getting severely infected by the mutated coronavirus. Lastly, unlike the first wave, the current spread of the virus mutants has reached remote locations (discussed later), where healthcare services are not sufficiently adequate. The purpose of this paper is to characterize the ongoing second wave so as to create awareness about the present grim situation and also sensitize the public as well as policymakers about the critical need for social distancing and other interventions to arrest further growth. We use several parameters such as testing rates, death rates as well as vaccination data in different states to describe the ongoing situation. Further, we also use available data to make informed projections of the epidemic growth based on a widely used epidemiological model.

## 2 Materials & Method

The present study is based on COVID-19 time-series data from the beginning of the pandemic to April 19, 2021. COVID-19 data for countries around the globe and Indian regional states are respectively taken from repositories maintained by *ourworldindata* (OWID; [8]) and *covid19india*.*org* (COVID19INDIA; [9]. OWID compiles data from the European Centre for Disease Prevention and Control (ECDC). COVID19INDIA curates data from several sources including the Indian government data, state bulletins, and official handles.

In order to identify the first and second waves, we study the *Effective Reproduction Number* (*R*_*t*_; [10]) as a marker for the decrease or surge in infections. *R*_*t*_ provides real-time feedback on the spread of pandemic as the *R*_*t*_ *>* 1 indicates a growth in infection, thus the goal is to implement social interventions to bring down *R*_*t*_ below 1 and close to 0 as much as possible. Time-varying *R*_*t*_ can be calculated using time series data of the infections and generation time distribution [11, 12]. We use the approach developed by Thompson *et al*. [13] for the estimation of effective Reproduction numbers using the R-package *EpiEstim*. A MATLAB implementation of this package, developed by Batista [14], is used for the current study. The generation time distribution requires serial interval as an input, which indicates the time between the onset of symptoms of a primary case and the onset of symptoms of secondary cases. Several studies [15, 16, 17, 18] have estimated serial interval for COVID-19. We take the mean serial interval as 4.7 days (95% *Confidence Interval* [CI]: 3.7, 6.0) with the standard deviation (SD) of the serial interval as 2.9 days (95% CI: 1.9, 4.9) based on Nishiura *et al*. [18].

The available dataset for the second wave has also been used for the predictions using the popular compartmental Susceptible-Infected-Recovered (SIR) model [19]. The incidence data between 27 February 2021 and 17 April 2021 have been used to obtain best-fit parameters used in the model. Note that the pandemic is still in the exponential-growth phase, as further expanded in the next section. Therefore, there is large uncertainty in the estimation of the peak of the daily cases, as well as the duration of the second wave. The details about the computation of underlying parameters and implementation of SIR model are given in [20, 21].

## 3 Results

We first analyze the available time-series data to quantify the virus transmissibility in India as well as in different states. Figure 3 exhibits the variations in daily confirmed cases as well as effective reproduction numbers (*R*_*t*_) in India. The effective reproduction number trend broadly follows the infection rate variation. In the first wave, the *R*_*t*_ value decreased from about 1.37 (95% CI 1.25 *−* 1.52) on 17 April 2020 to 1.09 (95% CI 1.07 *−* 1.11) on 10 September 2020. *R*_*t*_ went below the self-sustaining threshold of 1 for the first time on 23 September 2020 and remained there for the next 5 months, except for a minor flare up on 29 November for a couple of days. After this relatively long quiet interval, *R*_*t*_ started rising on 19 February 2021, which can be considered as an indicator of the arrival date of the second wave in India. The *R*_*t*_ has been increasing since then with minor fluctuations; as on April 19, 2021, *R*_*t*_ has reached approximately 1.37 (95% CI 1.28 *−* 1.47).

**Figure 3:**
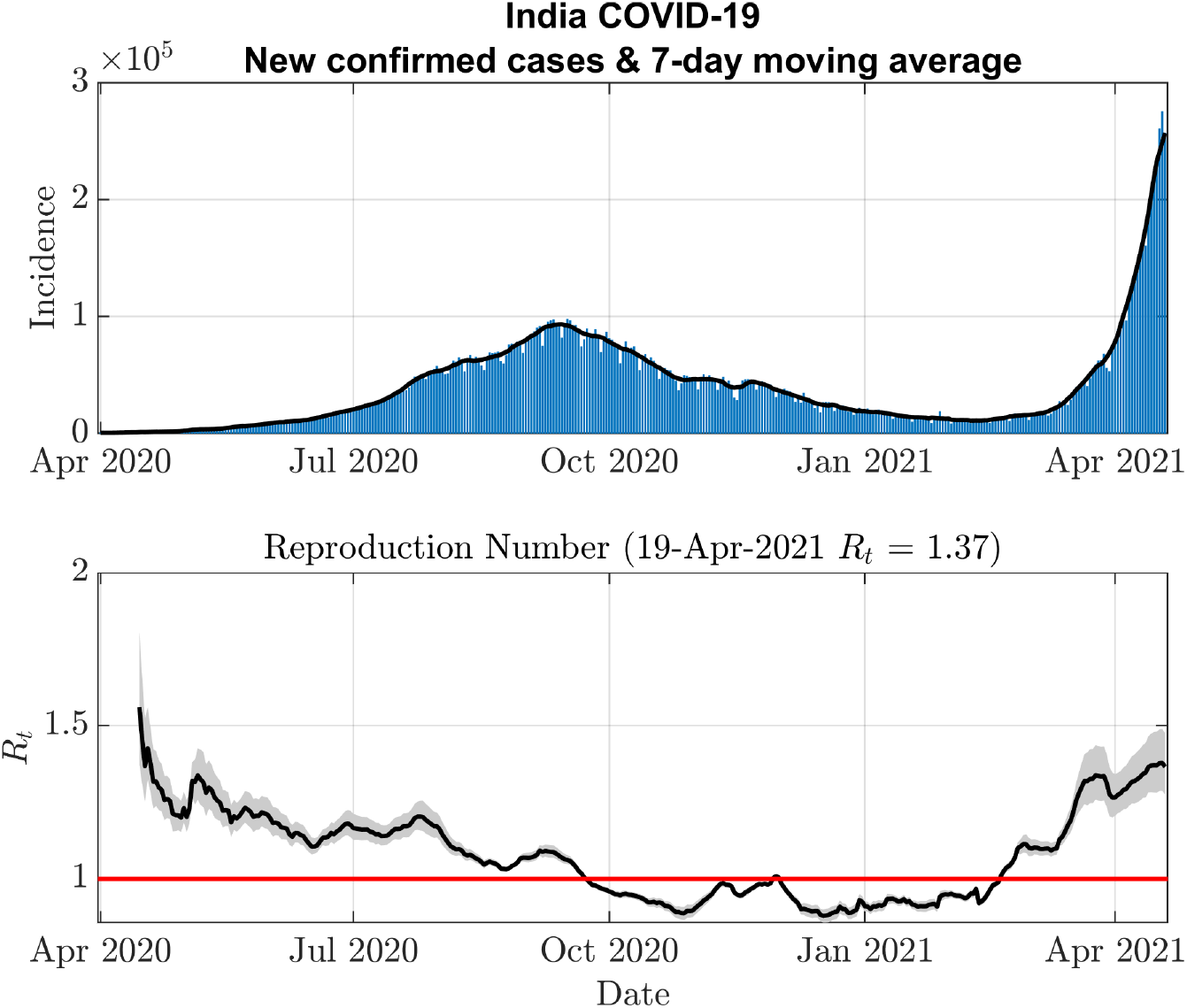
Temporal variations of the effective reproduction number (*R*_*t*_) along with incidence (daily infections) data. 95% CI of *R*_*t*_ variation is also shown in grey shadow. *R*_*t*_ was oscillating about 1.2 during the growth phase of the first wave and went below 1.0 for the first time around the end of September, 2020 marking the decline of the pandemic. *R*_*t*_ started rising again in February, 2021 and has a value of 1.37 on April 19, 2021.

Next, we describe the regional variations of effective reproduction numbers. Regional variations are expected to be dissimilar to one another commensurate with trends of new infections as shown in figure 2. Figure 4 exhibits these variations for the nine most-impacted states as of April 19, 2021, while the most recent values of *R*_*t*_ for all the states are reported in table 1. As evident in the figure, the incidence curves for these states show that the current daily cases are already higher than that of the first peak except for Andhra Pradesh. *R*_*t*_ curve crossed the threshold first in Maharashtra and about a week later in other states. However, very recently there has been a slight decline in the *R*_*t*_ value in Maharashtra (and also in Chhatisgarh and Punjab, which were highly impacted in the beginning of this wave). The most populated state (see last column in table 1), Uttar Pradesh, which was one of the least impacted states during the first wave, is currently in the high growth phase with *R*_*t*_ value of 1.74. Similarly, as shown in table 1, West Bengal and Bihar also have high *R*_*t*_ values. This is concerning as there is a large rural population in these states where the healthcare system is inadequate for such a scale of the pandemic.

**Figure 4:**
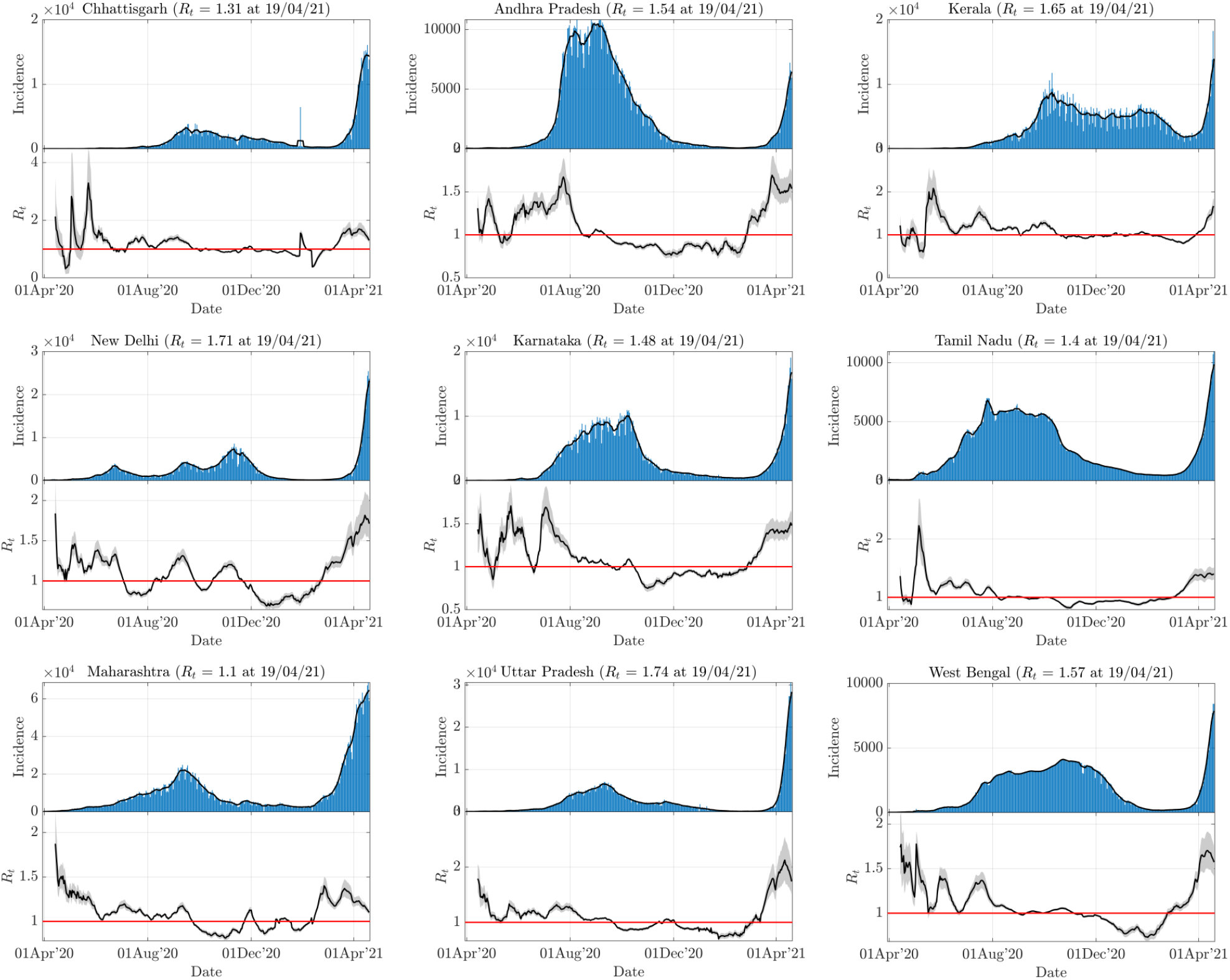
Temporal variations of the effective reproduction number (*R*_*t*_) along with incidence (daily infections) data for most-impacted states. Signs of the second wave were first noted in Maharashtra, where *R*_*t*_ first crossed the threshold of this year. All states have *R*_*t*_ *>* 1.0 as on April 19, 2021.

**Table 1:** Regional characteristics of the epidemic spread in Indian states and Union Territories as on April 19, 2021^*a*^.

| Region | CFR (%) | TPR (%) | CFR (%) | TPR (%) | $R_t$ | Vac. (%)<sup>b</sup> | Population |
| --- | --- | --- | --- | --- | --- | --- | --- |
|  | Cumulative |  | Daily |  |  |  | (in Crore) |
| Andhra Pradesh | 0.77 | 6.16 | 0.45 | 15.79 | 1.53 | 9.1 | 5.2 |
| Arunachal Pradesh | 0.33 | 4.05 | 0.00 | 5.59 | 2.52 | 12.0 | 0.15 |
| Assam | 0.51 | 2.86 | 0.51 | 2.09 | 1.88 | 4.9 | 3.4 |
| Bihar | 0.54 | 1.31 | 0.55 | 8.98 | 1.84 | 4.9 | 12 |
| Chandigarh | 1.21 | 9.45 | 0.65 | 20.36 | 1.21 | 12.7 | 0.118 |
| Chhattisgarh | 1.09 | 8.45 | 1.26 | 28.42 | 1.20 | 17.4 | 2.9 |
| Delhi | 1.41 | 5.38 | 1.01 | 26.12 | 1.72 | 13.4 | 2 |
| Goa | 1.32 | 11.46 | 1.81 | 34.48 | 1.43 | 16.5 | 0.154 |
| Gujarat | 1.32 | 2.58 | 1.03 | 7.20 | 1.48 | 15.6 | 6.8 |
| Himachal Pradesh | 1.52 | 5.61 | 0.77 | 30.04 | 1.36 | 10.9 | 2.9 |
| Haryana | 0.95 | 5.28 | 0.48 | 21.20 | 1.50 | 18.2 | 0.73 |
| Jammu and Kashmir | 1.39 | 2.18 | 0.40 | 4.45 | 1.35 | 13.0 | 1.3 |
| Jharkhand | 0.90 | 2.59 | 1.07 | 11.27 | 1.54 | 7.6 | 3.7 |
| Karnataka | 1.15 | 4.96 | 0.92 | 12.81 | 1.48 | 11.4 | 6.6 |
| Kerala | 0.40 | 8.73 | 0.15 | 15.63 | 1.65 | 17.2 | 3.5 |
| Madhya Pradesh | 1.10 | 5.91 | 0.61 | 25.32 | 1.53 | 9.2 | 8.2 |
| Maharashtra | 1.56 | 16.19 | 0.60 | 26.59 | 1.10 | 10.4 | 12.2 |
| Manipur | 1.27 | 5.01 | 1.85 | 5.58 | 1.93 | 5.1 | 0.31 |
| Meghalaya | 1.03 | 3.43 | 0.91 | 8.17 | 1.77 | 5.4 | 0.322 |
| Mizoram | 0.24 | 1.79 | 0.00 | 7.41 | 1.77 | 13.9 | 0.119 |
| Nagaland | 0.75 | 8.94 | 0.00 | 8.55 | 1.31 | 6.9 | 0.215 |
| Odisha | 0.54 | 3.87 | 0.09 | 12.29 | 1.80 | 11.5 | 4.4 |
| Puducherry | 1.48 | 6.55 | 0.88 | 15.85 | 1.4 | 11.2 | 0.15 |
| Punjab | 2.62 | 4.59 | 1.80 | 14.59 | 1.15 | 8.4 | 3 |
| Rajasthan | 0.75 | 5.47 | 0.44 | 29.78 | 1.63 | 14.5 | 7.7 |
| Sikkim | 2.03 | 8.01 | 0.00 | - | 1.74 | 25.2 | 0.066 |
| Tamil Nadu | 1.31 | 4.71 | 0.40 | 9.80 | 1.40 | 6.3 | 7.6 |
| Telangana | 0.52 | 3.01 | 0.35 | 4.82 | 1.48 | 8.5 | 3.7 |
| Tripura | 1.15 | 5.11 | 0.00 | 2.30 | 1.51 | 22.8 | 0.399 |
| Uttar Pradesh | 1.14 | 2.29 | 0.59 | 14.07 | 1.75 | 4.8 | 22.5 |
| Uttarakhand | 1.50 | 3.75 | 1.11 | 7.12 | 1.59 | 14.8 | 1.1 |
| West Bengal | 1.59 | 6.79 | 0.45 | 20.01 | 1.58 | 9.3 | 9.7 |

<sup>a</sup> Red-colored parameters indicate greater probability of virus spread<sup>b</sup> At least one dose is administered

For perspective, we compare the *R*_*t*_ value of India with those of other most impacted countries as on April 19, 2021 in table 2. Except for the UK, where the pandemic is in decline (*R*_*t*_ ≃ 0.90), the other four countries have *R*_*t*_ close to the threshold value. The fraction of the vaccinated population is also listed in the table. Among all countries, the US and UK have the highest vaccinations per 100 persons; these countries are possibly least susceptible to another wave provided the vaccines can provide protection against change in virus phenotype due to mutations [22]. India, despite running one of the largest vaccination efforts in the world [7] with two made-in-India vaccines – Covaxin and Covishield, has a relatively low share of vaccinated people per capita due to a very large population (see last column in table 2). Further, the population covered so far is above 45 years of age except for healthcare professionals and frontline workers. The vaccination drive needs to be enhanced to include the younger population, say from 18 years of age, as several reports suggest that mutated virus may have higher virulence towards younger population than that in the first wave [23]. As of April 19, 2021, India has approved other vaccines to be procured and administered, which could further accelerate the vaccination effort.

**Table 2:** Characteristics of the epidemic spread in six most-affected countries as on April 19, 2021.

| Country | CFR (%) | TPR (%) | CFR (%) | TPR (%) | $R_t$ | Vac. (%) | Population<br>(in Crore) |
| --- | --- | --- | --- | --- | --- | --- | --- |
|  | Cumulative |  | Daily |  |  |  |  |
| US | 1.79 | 7.54 | 0.94 | 3.18 | 1.01 | 63.3 | 33.2 |
| India | 1.18 | 5.72 | 0.68 | 15.65 | 1.37 | 9.5 | 139.1 |
| Brazil | 2.68 | - | 4.64 | - | 1.012 | 15.9 | 21.4 |
| France | 1.91 | 7.29 | 1.49 | 7.11 | 0.93 | 25.1 | 6.5 |
| Russia | 2.25 | 3.74 | 4.03 | 1.43 | 0.99 | 11.1 | 14.6 |
| UK | 2.90 | 3.05 | 1.0 | 0.18 | 0.90 | 63.5 | 6.8 |

To further characterize the second wave, we employ the following well-known ratios:

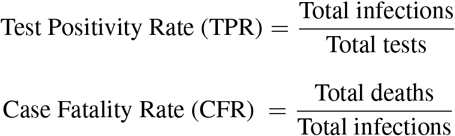

These ratios can be calculated based on the cumulative data or daily data. While the estimates based on cumulative data are more smooth, the daily ratios reflect sudden changes more prominently. Therefore, we employ both definitions to illustrate different aspects, although the daily ratios are obtained after a 7-day moving average to remove large fluctuations due to reporting delays and other uncertainties.

The Test positivity rate (TPR) typically indicates whether the number of tests is enough to contain the spread of the virus by isolating and quarantining the positive cases. Figure 5(a) illustrates the temporal variations of these parameters based on cumulative data for both first and second waves. India shows an increase in cumulative TPR during the acceleration phase of the first wave and it starts declining from August 2020. Recently, the TPR curve shows an upward trend commensurate with the spurt of cases, with a TPR value of 5.72% as on April 19, 2021. This increase in TPR in the second wave is better reflected in the daily estimates as shown in figure 5(b). The daily TPR curve shows a sharp spike in its value since late March 2021. As on April 19, 2021, the daily TPR value for India is about 15%. WHO recommends that this TPR value should be less than 5% for at least two weeks so that the transmission can be brought under control. A sudden increase in daily TPR to a very high value indicates an alarming situation and necessitates ramping up of daily testing capacity.

**Figure 5:**
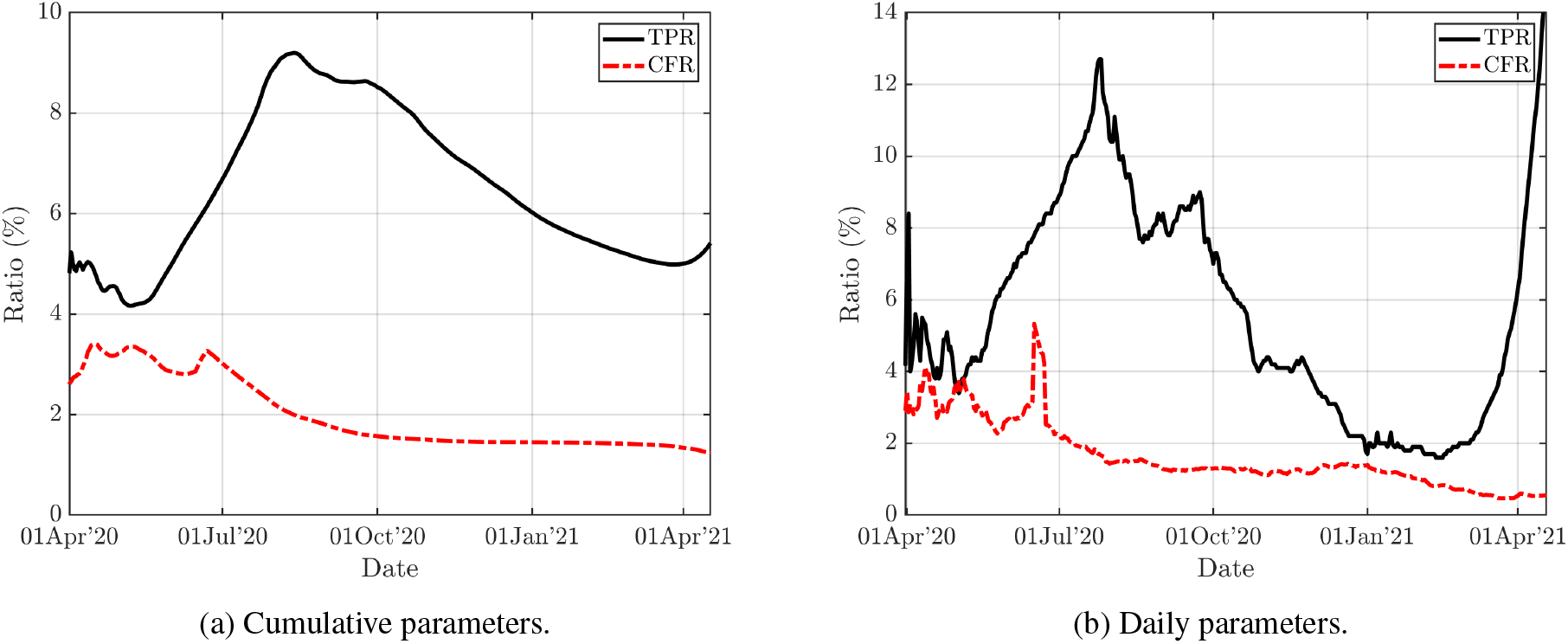
Temporal variations of two characterizing parameters-Test positivity rate (TPR) and Case fatality rate (CFR)- are shown. These ratios are computed using the cumulative data in (a) and daily data in (b). There is a sudden increase in TPR in the second wave which presents a worrying situation. CFR in the second wave is lower than that in the first wave. However, (b) shows signs of its increase very recently. This may be due to reduced healthcare access to the patients due to a drastic increase in the number of infections.

Further, there are large variations among different regions in India as shown in Table 1. Ten states have alarming daily TPR of more than 20% as highlighted in red. Maharashtra, which shows a slight decline in *R*_*t*_ value recently, has a very high TPR of 26.6% indicating very high transmission. Therefore, the actual number of infections is likely to be higher than that being reported due to a limited diagnostics capacity. States which have a high *R*_*t*_ value, as well as high daily TPR (*>* 10%), are also at significant risk.

Next, we report the case fatality rate for India in figures 5 (a) and (b) based on cumulative and daily data respectively. Both the CFR curves show downward trends with time. The cumulative CFR curve goes from 3.5% in mid-April 2020 to 1.2% in mid-April 2021 with minor fluctuations. Interestingly, although the second wave shows the virus to be more infectious, the decline in the CFR curve suggests a silver lining of a relatively less fatal mutant. However, considering an exponential increase of cases at a very high rate, it is expected that soon the healthcare facilities will be fully throttled resulting in the unavailability of hospital beds and ventilators to those in critical needs. This may result in an increase in CFR. Even otherwise, in terms of absolute numbers, the daily deaths are already higher than the level of peak values in the first wave (see figure 1(b)). Table 1 lists statewise data of cumulative and daily CFRs, with Punjab having the highest value and Kerala the least among severely-affected states.

Finally we describe the vaccination data in states as listed in Table 1. Among the most-impacted states with sizeable population, Kerala and Chhatishgarh have highest vaccination per 100 people. Uttar Pradesh and Bihar, which have very high *R*_*t*_, have the lowest level of vaccination per capita. This further suggests the need for strong interventions in the these states while vaccination capacity is increased simultaneously with prioritized allocations based on social contacts [24].

After the characterization of the second wave, we employ mathematical and epidemiological models to understand the dynamics and to provide actionable insights. As discussed earlier (see figure 2), the second wave started slowly near 11 February 2021 in Maharashtra and Punjab, but the epidemic spread quite rapidly in other states from early March. Therefore we use two exponential curves to fit the incidence (daily cases) data. Further, we compare this fit with that of the first wave, starting from a date when the number of infections was similar to that in the second wave. Figure 6 exhibits these exponential fits, while the coefficients, as well as statistical fitness data, are listed in Table 3. The first exponential fit of the second wave has a lower exponent than the first wave (compare *b*_1_ and *b*_2_ in table 3), although it has a relatively low value of the coefficient of determination (adjusted *R*^2^ ≃ 0.62).

**Figure 6:**
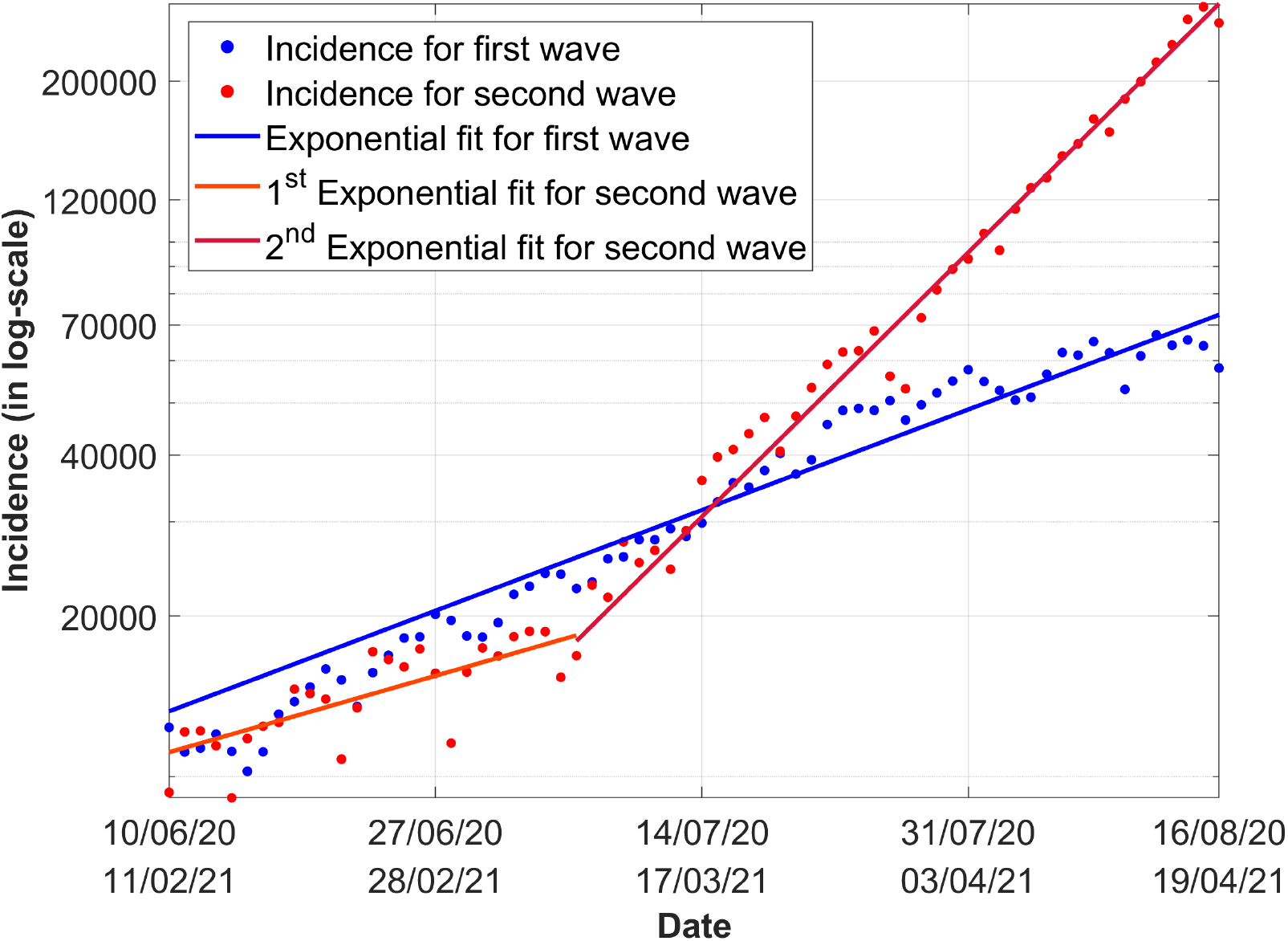
Comparison between first and second waves in equal interval of time starting from dates when the number of infections were almost similar. In the first wave, data between 10 June 2020 and 16 August 2020 are considered, which shows an exponential fit. In the second wave, two different exponential fits are used corresponding to green and orange regions shown in figure 2. The exponential growth factor in the second fit on the recent data is more than double in that of the first wave fit.

**Table 3:** Exponential regression model for initial growth in first and second waves: *y* = *a* exp(*bt*).

| Wave | Coefficients | Estimate | 95% CI | | SE | tStat | pValue | adjusted $R^2$ |
| --- | --- | --- | --- | --- | --- | --- | --- | --- |
| First | $a_1$ | $1.22 \times 10^4$ | $1.12 \times 10^4$ | $1.33 \times 10^4$ | 531 | 23.0 | $9.79 \times 10^{-32}$ | 0.955 |
| | $b_1$ | 0.0278 | 0.0261 | 0.0296 | 0.000896 | 31.0 | $4.36 \times 10^{-39}$ | |
| Second(1) | $a_2$ | $1.11 \times 10^4$ | $0.99 \times 10^4$ | $1.23 \times 10^4$ | 578 | 19.2 | $1.72 \times 10^{-16}$ | 0.616 |
| | $b_2$ | 0.0194 | 0.0131 | 0.0256 | 0.00305 | 6.35 | $1.18 \times 10^{-06}$ | |
| Second(2) | $a_3$ | $2.65 \times 10^3$ | $2.23 \times 10^3$ | $3.07 \times 10^3$ | 208 | 12.75 | $1.13 \times 10^{-15}$ | 0.991 |
| | $b_3$ | 0.0689 | 0.0662 | 0.0715 | 0.00129 | 53.3 | $8.77 \times 10^{-39}$ | |

However, the exponent of the second fit on more recent data in the second wave is more than twice of the first wave that explains the rapid growth of the pandemic. Fits for both the first wave and rapid second wave are statistically significant with adjusted *R*^2^ greater than 0.95.

We now employ the dynamical SIR model [25] for an estimate of the progression of the pandemic. This model takes both the incidence and recovery data to estimate the peak as well as the eventual decline of the pandemic. SIR model is a compartmental model, where the total population is divided into three segments-susceptible, infected, and recovered (sometimes also termed as removed). Using the parameters - infection (*β*) and recovery (*γ*) rates, the model plots the progression of disease with time in terms of these three segments using simple governing equations. This model has been widely used for the prediction of COVID-19 in several countries [26, 27]. More sophisticated variations of these models that further subdivides the population into ‘exposed’ (SEIR; [28]), ‘asymptomatic’ (SAIR; [29]), ‘exposed-quarantined-deceased-insusceptible’ (SEIQRDP; [30]) are also used for predictions of the first wave in India. A review of the predictive models can be found in [31].

Note that forecasting the peak (both the time as well as numbers) with any model is difficult when the pandemic is in the exponential growth phase. This is because there is large uncertainty in the estimation of underlying parameters for these models using the infection data only from the exponential growth phase. Pandemics usually follow power-law beyond the exponential growth [32, 33], during which the estimation of the peak is more accurate. Nonetheless, the projections provide ballpark estimates and therefore help policymakers to prepare for the future and plan necessary actions. Figure 7 shows the predictions using the SIR model with parameters obtained from the data between 27 February 2021 and 17 April 2021. As shown, the peak of the second wave is estimated in the middle of May with the peak number of daily cases of more than 0.4 million. Furthermore, even with a low daily CFR of 0.68% as of April 19, 2021, India will have about 3,000 deaths per day at its peak. This number is expected to increase as access to healthcare becomes more and more limited with the surge in active infections.

**Figure 7:**
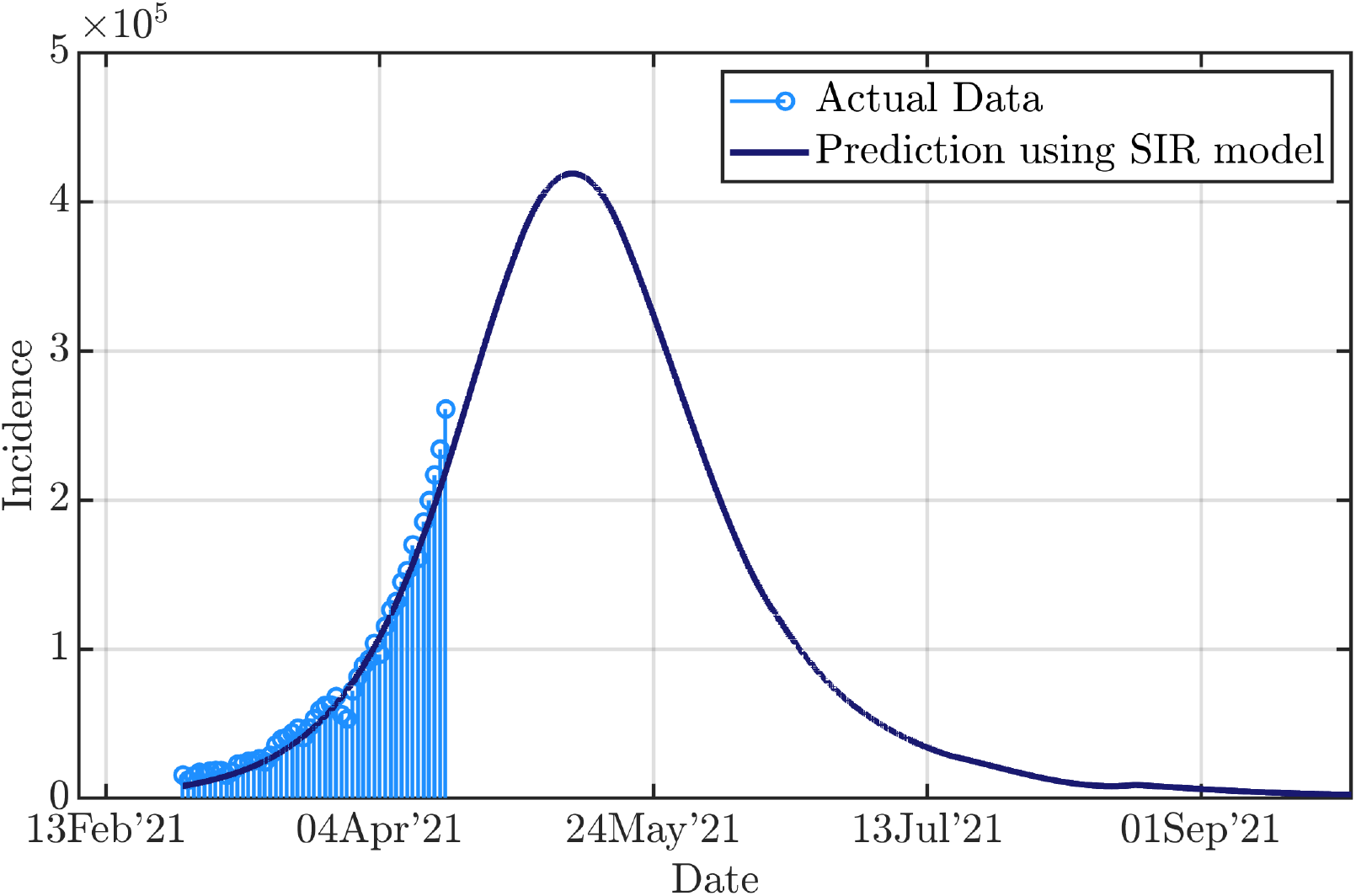
Predictions of the COVID-19 epidemic in India using classical Susceptible-Infected-Recovered (SIR model). Data between 27 February 2021 to 17 April 2021 are taken for fitting of parameters. The predictions suggest the second wave to peak around mid-May with infections exceeding 0.4 million. Note that uncertainty in infection data due to high positivity rate and regional variations in the spread of virus as well as interventions are not taken into account for predictions. Consideration of these factors could alter the estimates.

An important point to note here is that as mentioned above the high TPR of India suggests that the actual number of infections could be much higher than the reported infections. Since the model parameters are estimated using the reported number of infections, the projection of the peak case count is still conservative. Therefore, the results presented above should be taken as an optimistic scenario, provided no new interventions are introduced. The high TPR also suggests that India may see a plateau (of about 0.4 million daily for several days) in May instead of a sharp peak as testing capacity limits the number of daily cases. Thus, while the growth rate may be negligible in this period, active cases will continue to pile up therefore throttling the healthcare infrastructure. Note that the current model does not include several factors such as asymptomatic cases, partial or full lockdowns, vaccination data, regional heterogeneities etc. These factors may be considered for more realistic modelling of the second wave once more data is available. We plan to employ universal curve [34] and refined epidemiological models [26, 35] for improved prediction of the second wave in India.

## 4 Conclusions

The second COVID-19 wave in India, which began in the second week of February, 2021, presents a grim situation with the number of daily cases reaching around 0.3 million on April 19, 2021. The data suggests that at present the virus is much more infectious than the first wave, but the number of daily deaths per infection is lower. However, with an inordinate increase in the number of cases and over-stretched healthcare system, the daily death count may increase substantially. The effective reproduction number (*R*_*t*_) is estimated for India, as well as for the Indian states. At present, almost every state show *R*_*t*_ *>* 1 suggesting that the second wave has spread everywhere including rural areas which were largely untouched during the first wave. This includes populous states like Uttar Pradesh, Bihar, and West Bengal: each has *R*_*t*_ value greater than 1.58. With rural areas impacted, it may be necessary to take aggressive lockdown measures to arrest the further spread while sufficient amount of vaccine becomes available. The SIR model based on available data suggests the peak of the epidemic to occur in the middle of May 2021 with approximate daily infections exceeding 0.4 million.

In summary, using the available infection data, we analyze the second COVID-19 wave in India. We observe that the epidemic is creating unprecedented havoc in the population. We hope that the appropriate administrative intervention, aggressive vaccination drive, and people’s participation will help flatten the curve earlier than the grim forecast of the epidemic.

## Data Availability

The data included in the study are publicly available.

https://ourworldindata.org/

https://www.covid19india.org/

## 5 Acknowledgment

This material is based partly upon work supported by a SERB MATRICS project SERB/F/847/2020-2021.

